# Psychophysical Olfactory Findings of Mild-to-moderate COVID-19 Patients: Preliminary Report

**DOI:** 10.1101/2020.05.02.20070581

**Authors:** Jerome R. Lechien, Pierre Cabaraux, Carlos M. Chiesa-Estomba, Mohamad Khalife, Stéphane Hans, Delphine Martiny, Sven Saussez

## Abstract

Since the onset of the COVID-19 infection, many patients reported sudden loss of smell (SLS). However, due to the lack of psychophysical testings, it remains difficult to know if these patients really have hyposmia or anosmia. Our group investigated the prevalence of anosmia and hyposmia in 28 COVID-19 patients and the potential association with nasal complaints.

## Methods

Adults with SLS and mild-to-moderate COVID-19 manifestation were recruited after a public call of the University of Mons (Belgium). The disease was confirmed through nasopharyngeal swab (RT-PCR). Patients with neurological disorder, chronic rhinosinusitis or history of nasal surgery were excluded. The patient epidemiological characteristics and the otolaryngological symptoms were electronically collected through an online questionnaire (Professional Survey Monkey®, San Mateo, California, USA). The nasosinusal symptoms were electronically evaluated through the French sino-nasal outcome test-22 (SNOT-22).^2^

The olfactory and gustatory questions were based on the smell and taste component of the National Health and Nutrition Examination Survey.^3^ The questions were chosen to characterize the variation, timing and associated-symptoms of olfactory and gustatory dysfunctions. The impact of olfactory dysfunction on quality of life of patients was evaluated with the short version of Questionnaire of Olfactory Disorders-Negative Statements (sQOD-NS).^3^

Patients benefited from psychophysical olfactory evaluation through sniffin stick tests (Medisense, Groningen, Netherlands): 16 pens were presented to patients every 30 seconds. The patient has to choose the adequate term describing the smell between 4 given options. The test was scored on a total of 16 points and allowed categorization into in 3 groups: normosmia (score between 12-16), hyposmia (score between 9-11) and anosmia (score <9).

The relationship between clinical and olfactory outcomes was analyzed through multiple linear regression (SPSS, v22,0; IBM-Corp, Armonk, NY, USA).

## Results

28 patients completed the study (19 females). At the time of the evaluation, the mean duration of olfactory disorders was 10.8±3.8 days (Table 1). The most common otolaryngological symptoms were rhinorrhea, postnasal drip and nasal obstruction. Aroma disorder and dysgeusia, which was defined as the impairment of the following taste modalities: salty, sweet, bitter and sour, accounted for 85.7% and 60.1% of cases. Cacosmia and phantosmia occurred in 46.4% and 25% of patients, respectively.

**Table 1.** Epidemiological & Clinical Characteristics of Patients.

footnotes: Abbreviations: GERD=gastroesophageal reflux disease; SD=standard deviation; SNOT-22= sino-nasal outcome-22 questionnaire; sQOD-NS: short version of Questionnaire of Olfactory Disorders-Negative Statements.
| Characteristic | All patients<br>(N=28) |
| --- | --- |
| Age |  |
| Mean (SD) - yo | 44.0 ± 12.0 |
| Gender (N - %) |  |
| Female | 19 (67.9) |
| Male | 9 (32.1) |
| Ethnicity (N - %) |  |
| Caucasian | 26 (92.9) |
| North African | 2 (7.1) |
| Addictions (N - %) |  |
| Non-smoker | 24 (85.7) |
| Mild smoker (1-10 cigarettes daily) | 4 (14.3) |
| Moderate smoker (11-20 cigarettes daily) | 0 (0) |
| Heavy smoker (>20 cigarettes daily) | 0 (0) |
| Allergic patients | 5 (17.9) |
| Comorbidities |  |
| Hypertension | 4 (14.3) |
| Allergic rhinitis | 3 (10.7) |
| GERD | 3 (10.7) |
| Asthma | 2 (7.1) |
| Hypothyroidism | 1 (3.6) |
| Depression | 0 (0) |
| Autoimmune disease | 0 (0) |
| Sarcoidosis | 0 (0) |
| Hemochromatosis | 0 (0) |
| Psoriasis | 0 (0) |
| Obstructive apnea syndrome | 0 (0) |
| Diabetes | 0 (0) |
| Renal failure | 0 (0) |
| Hepatic insufficiency | 0 (0) |
| Respiratory insufficiency | 0 (0) |
| Heart problems | 0 (0) |
| Ear, Nose & Throat Symptoms Prevalence |  |
| Nasal obstruction | 15 (53.6) |
| Rhinorrhea | 15 (53.6) |
| Postnasal drip | 15 (53.6) |
| Throat pain | 9 (32.1) |
| Facial pain | 7 (25.0) |
| Ear pain | 10 (35.7) |
| Dyspnea | 5 (17.9) |
| Dysphagia | 5 (17.9) |
| Dysphonia | 7 (25.0) |
| Dysgeusia (salty, sweet, bitter & sour) | 17 (60.1) |
| Aroma Disorders (sense reduction or distortion) | 24 (85.7) |
| Duration of dyspnea, days (mean, SD) | 10.8 ± 3.8 |
| SNOT-22 Total score | 37.3 ± 17.0 |
| sQOD-NS | 14.8 ± 4.4 |

The psychophysical olfactory evaluations reported that 53.6% and 21.4% of SLS patients were anosmic and hyposmic, respectively; 28.6% being normosmic (N=8). Among the 15 anosmic patients, 7 (46.7%) did not report nasal obstruction, rhinorrhea or postnasal drip during the disease clinical course. Among the 6 hyposmic patients, only one did not report these complaints. There were no significant differences between groups about the SLS mean duration.

Patients with stronger sniffin test results had stronger sQOD-NS score (p=0.036). Anosmic patients more frequently reported dyspnea, gustatory disorder and face pain (p<0.020). There was no significant association between the results of the sniffin tests, the SNOT-22 items/total score and the following complaints: nasal obstruction, rhinorrhea and postnasal drip.

## Discussion

The olfactory disorder seems to be a key symptom of mild-to-moderate European and American COVID-19 patients, accounting for more than 70% of patients.^1,4^ However, our preliminary results suggested that only 53.6% of patients who claimed that they have SLS really have anosmia regarding psychophysical tests. Regarding to this mismatch, the prevalence of olfactory dysfunction related to COVID-19 would be overestimated in the epidemiological studies based on patient-reported outcome questionnaires.

The majority of anosmic had no nasal complaints, which are usually associated with viral infection, nasal mucosa inflammation and related-SLS. These data may support the complexity of the mechanisms underlying the development of SLS in COVID-19. As supported by basic science researches, SLS would be related to neural spread of the virus through the olfactory cleft mucosa, affecting the olfactory neurons or glial cells.^5^ The neurotropism of some coronavirus strains had previously been demonstated.^6^ Future studies are needed to confirm the data of the current paper.

## Data Availability

All data are available for consultation by asking

## Author contribution

*Study concept and design:* Lechien, Cabaraux, Chiesa, Saussez, Hans.

*Acquisition, analysis, or interpretation of data*: Cabaraux, Lechien, Saussez, Khalife, Martiny. *Drafting of the manuscript:* Lechien, Saussez.

*Critical revision of the manuscript for important intellectual content:* Saussez, Chiesa, Hans.

## Acknowledgments

Dr Hopkins for their relevant comments.

